# Physical body experiences questionnaire simplified for active aging (PBE-QAG): Validation with Rasch measurement theory

**DOI:** 10.1101/2021.06.01.21258192

**Authors:** Wei Deng, Sydney Carpentier, Ann Van de Winckel

## Abstract

**Purpose:** To validate the Physical body experiences questionnaire simplified for active aging (PBE-QAG) with Rasch measurement theory. PBE-QAG measures body awareness during physical activity and includes dimensions of body-mind relationship, body acceptance, and awareness of physical skills and limits.

**Methods:** Adults without pain (n=269), with pain (n=61), with mental health conditions (n=200), and with stroke (n=36) were recruited at the Minnesota State Fair, Highland Fest, and in the Brain Body Mind Lab (University of Minnesota) and completed demographic and clinical questionnaires as well as the PBE-QAG. The PBE-QAG has 12 items, with scores ranging between 0 (totally true) to 4 (totally false). A low total score on the PBE-QAG reflects better body awareness. We evaluated item and person fit, targeting, unidimensionality, person separation reliability (PSR), local item dependence (LID), and differential item functioning (DIF) for demographic and clinical characteristics. We compared with Kruskal-Wallis ANOVA the person mean location in four groups: Adults with or without mental health conditions; and whether those groups did body awareness training.

**Results:** Unidimensionality and item fit were obtained after deleting 2 and rescoring 5 items. Seven participants did not fit the model (1.23%). There was minimal floor (5.72%), no ceiling effect (0.00%), and no LID. No DIF was greater than 0.50 logits for any of the variables. The Wright-corrected PSR was 0.96. The person mean location was -1.71±1.21 logits. Adults with mental health conditions who did not practice body awareness had a higher person mean location [Median (IQR)=0.83(0.89) logits, *p*<0.0001] versus the other three groups, reflecting lower body awareness.

**Conclusions:** PBE-QAG demonstrated good item and person fit, but the targeting is off. Therefore, the current version of PBE-QAG is not recommended for use in the general population. We encourage further validation of PBE-QAG in adults with mental health conditions who do not practice body awareness.

## Introduction

Adults with altered body awareness –such as adults with mental health conditions (i.e., anxiety and depression), with chronic pain, or with stroke– can improve their physical and mental functioning by practicing mindfulness or a mind and body approach such as yoga, Tai Chi, or Qigong [1–7]. Such practices help restore physical and mental health through at least two mechanisms: First, dysfunction in the autonomic nervous system is closely implicated in the onset and maintenance of disease states, and mind and body approaches help rebalance this dysfunction [8,9]. Second, these practices improve body awareness because they require a focused attention on the present moment, on bodily sensations, movements, thoughts, or feelings, and on stimuli from the environment, with a non-judgmental awareness and with acceptance [10–13]. When adults are aware of their body in the present moment, they can also more easily notice physical or mental unease and thus take action to regain a healthier state [13].

Body awareness in the context of mind and body approaches is defined as a perceptual understanding and awareness of (i) proprioception: body position and movement sense and how those body parts are situated in peripersonal space, (ii) exteroception: visual, tactile, auditory, and pain signals and their locations within the peripersonal space; and (iii) interoception: internal body states [13–21]. In addition, yoga training as well as regular physical training have been associated with adults having a more positive relationship with their body [22,23], and having a more conscious awareness of their physical skills and limitations [24].

Yet, because body awareness is a multidimensional concept, clinicians and researchers need reliable and valid assessments to measure changes in well-defined aspects of body awareness in their patients or participants, after they have completed mind and body treatments or a physical activity program. Therefore, these assessments should measure those relevant aspects of body awareness that adults with mental health conditions, with pain, or with stroke may have deficiencies in. Furthermore, it is necessary to know what the “normal scoring ranges” of those aspects of body awareness are, so that clinicians can properly interpret changes or improvements in patient populations.

The “Physical Body Experiences Questionnaire simplified for active aging (PBE-QAG)”[25] is a 12-item scale that was inspired by a previously described instrument [26], but modified, simplified, and adapted for adults over 65 years of age. This questionnaire covers statements about four dimensions of body awareness related to physical activity (i.e., body-mind relationship, body acceptance, awareness of physical skills, and awareness of physical limitations). Those four dimensions were identified in a previously cited study in older adults with depression [27]. Participants respond to which extent those statements are true for them, by choosing a response between 1 (totally true) to 5 (totally false), with lower scores representing a higher level of body awareness.

Both Menzel (2010) and Cossu *et al*. (2018) confirmed the existence of the above mentioned four dimensions through confirmatory factor analysis and reported acceptable to good internal consistency for each of those domains (Cronbach’s alpha between 0.61-0.83) [25,26]. Cossu *et al*. (2018) further demonstrated that people who did more than 10 min of physical activity per week had a lower score on the PBE-QAG, reflecting a higher level of body awareness related to physical activity [25]. While those analyses give some indication on dimensionality (confirmatory factor analyses), and internal reliability (Cronbach’s alpha), they are insufficient to encompass the psychometric rigor needed to promote a scale for use in the clinic or in research. Rasch Measurement Theory transforms an ordinal scale into an interval scale level of measurement, by accounting for both the participant’s level of ability on the investigated trait and the relative item difficulty [28,29].

Therefore, as a next step in obtaining a more rigorous psychometric analysis of the PBE-QAG, the aims of this study were to evaluate the structural validity of the PBE-QAG with Rasch Measurement Theory in (i) healthy adults, to know which score range would be considered ‘normal’ body awareness related to physical activity; and in (ii) adults with known body awareness deficits, such as adults with mental health conditions, with pain, or with stroke, to verify whether this scale can accurately measure body awareness deficits in these populations.

## Materials and methods

### Participants

We recruited adults in the community and adults with chronic stroke between the ages of 18-99 years through volunteer sampling by means of flyers and website announcements. Additionally, adults signed up for research by expressing their interest at research booths at the Minnesota State Fair and Highland Fest. We also reached out per email to a list of healthy volunteers who had contacted the Brain Body Mind Lab (University of Minnesota) to express their interest in participating in research. We excluded pregnant women and adults who did not speak English. The study was approved by the University of Minnesota’s Institutional Review Board (IRB# STUDY00005849 and STUDY00000821) and the study was performed in accordance with the Declaration of Helsinki.

The healthy adults at the State Fair and Highland Fest, as well as healthy adults who received an email, accessed the REDCap questionnaires through a secure REDCap web link. After reading and providing consent, they were quizzed on the comprehension of the content of the consent form through the University of California, San Diego Brief Assessment of Capacity to Consent (UBACC) [30]. They then completed the survey on a tablet (at the State Fair or Highland Fest) or on their own computer. All information was stored automatically on the secure University of Minnesota REDCap platform. We did not collect any personal or identifiable information from healthy adults and from any adults signing up for research at the Minnesota State Fair or Highland Fest. Adults with stroke signed informed consent on paper and their assessments were collected on paper.

### Main outcome measures

The survey encompassed questions on general health, which included questions on pain, mental health conditions, or presence of conditions or diseases, as well as questions about demographics and current practices of mindfulness, breathing exercises or body awareness training, such as yoga, Qigong, or Pilates.

As mentioned in the introduction, the PBE-QAG measures four dimensional factors of body awareness related to physical activity (i.e., mind-body relationship, body acceptance, awareness of physical skills, and awareness of physical limits), with a lower total score on the items representing a higher level of body awareness related to physical activity. In order to perform the Rasch analysis, we reorganized the scoring categories 1-5 to scoring categories 0-4.

## Statistical analysis

Rasch Measurement Theory is based on a probabilistic model, which states that a person with higher ability on a certain trait should have a higher probability of obtaining a better score (in our case, a better score is a lower score). Rasch analysis transforms an ordinal scale into an interval scale with the ability of the persons and difficulty of the items represented on the same log-odds ratio (logits) units. We used Rasch Unidimensional Measurement Model (RUMM) 2030 software (RUMM Laboratory, Perth, WA, Australia) for the Rasch analysis.

To assess structural validity and unidimensionality, we evaluated the presence of disordered thresholds, item and person fit, person separation reliability (PSR), local item dependence (LID), differential item functioning (DIF), principal components analysis of residuals (PCAR), and targeting, the latter which includes floor-and ceiling effect. A scale is considered unidimensional when all items reflect the same underlying construct. Disorder thresholds of items indicate that the order of the response categories is not following the logical estimated order of the underlying construct [31,32]. Individual person-and item-fit statistics are reported as Residuals and Chi-square statistics, with Fit Residuals greater than +2.50 and less than -2.50 indicating item misfit, and item redundancy, respectively.

The PSR is a measure of precision that reflects how well persons can be differentiated into different levels of low and high ability of the trait in order to serve the clinical or research purpose [33]. To distinguish different ability levels among persons, this value should be at minimum 0.70 for group decisions; and over 0.90 in order to make decisions for individuals [28,34,35]. Another measurement of precision is the mean error variance, reflecting the Root Mean Square standard Error computed over the persons or over the items [36]. Floor and ceiling effects become problematic when 15% or more of the sample obtains a minimum or maximum score on the assessment [37]. Adequate targeting of the assessment is obtained when the average person location is within a range of -0.50 and 0.50 logits of the average item location [38].

DIF evaluates whether the hierarchy of the items of the measurement is preserved across demographic or clinical factors [39]. A shift greater than 0.50 logits is considered evidence of DIF [40]. In this study, we evaluated whether the following demographic or clinical parameters would differentially affect the item structure: sex (female, male, other); age (below 65 or above 65 years of age); the presence of pain (yes, no); groups (healthy adults, adults with mental health conditions, adults with pain, adults with stroke); currently having a breathing practice (yes, no); currently doing relaxation exercises such as mindfulness exercise, meditation or body scan exercises (yes, no); and currently doing any type of body awareness training such as Tai Chi, Qigong, martial arts, yoga, dance, etc. (yes, no).

PCAR indicates the extent to which variance in the residuals is random. If so, then unidimensionality is implied because this variance is then explained by the underlying trait, and not by other underlying traits. PCAR’s output of the first contrast will reflect this with an eigenvalue of less than 2 and with the percent variance explained by this first contrast being less than 10% [41]. If this is not the case, then unidimensionality can be further tested by performing a series of independent *t*-tests of 2 subtests of items that load respectively positively and negatively (correlations above and below 0.30) on the first contrast. Multidimensionality is noted when more than 5% of these *t*-tests are significant [28].

Finally, LID occurs when a pair of items shares a greater degree of content with each other than with other items of the assessment. LID is reported for standardized residual item correlations that are at least 0.20 above the average residual item correlation.[42] For all analyses, where appropriate, Bonferroni corrected *p*-values are reported.

## Results

### Demographic and clinical data

We recruited 566 adults among which 269 were healthy adults (age=51±17 years), 200 were adults with self-reported mental health conditions (age=47±16 years), 61 were adults with self-reported pain (age=58±14 years), and 36 were adults with chronic stroke resulting in left or right hemiplegia or hemiparesis of the upper limb (age=58±13 years). Table 1 displays the demographic and clinical data of each of those groups.

**Table 1.** Demographic and clinical characteristic of participants by group.

|  | Healthy<br>adults<br>(n=269) | Adults with<br>mental health<br>conditions<br>(n=200) | Adults with<br>pain<br>(n=61) | Adults with<br>chronic<br>stroke<br>(n=36) |
| --- | --- | --- | --- | --- |
| Age (years, mean±SD) | 50.91±17.01 | 46.66±16.41 | 57.72±13.85 | 58.00±12.93 |
| Gender |  |  |  |  |
| Female | 113 | 52 | 26 | 11 |
| Male | 155 | 148 | 35 | 25 |
| Other | 1 | 0 | 0 | 0 |
| Level of education (years, mean±SD) | 5.84±1.77 | 5.85±1.93 | 5.89±1.84 | 5.56±1.66 |
| Ethnicity |  |  |  |  |
| Not Hispanic or Latino | 266 | 196 | 60 | 36 |
| Hispanic or Latino | 3 | 4 | 1 | 0 |
| Racial background |  |  |  |  |
| American Indian or Alaska Native | 1 | 0 | 1 | 0 |
| Asian | 20 | 7 | 3 | 2 |
| Black/ African American | 5 | 5 | 1 | 1 |
| White | 234 | 180 | 53 | 30 |
| Multiracial | 4 | 5 | 1 | 2 |
| Other | 5 | 3 | 2 | 1 |
| Pain | 0 | 47 | 61 | 18 |
| Mental health conditions | 0 | 200 | 0 | 2 |
| Current breathing exercise | 95 | 100 | 22 | 13 |
| Current relaxation exercise | 72 | 87 | 13 | NA |
| Current body awareness training | 82 | 72 | 16 | 13 |

**Legend:** NA = not assessed

### Rasch Measurement Theory

The iteration analysis table (S1 Table) shows the step-by-step iteration of the Rasch analysis performed on the PBE-QAG. Per iteration step, the table displays the number of rating scales categories and number of items, person mean location in logits, mean error variance, floor and ceiling effect, overall fit, item and person fit, number of items with disordered thresholds, PCAR, and PSR at every iteration step. We provided a Wright-corrected PSR (PSRw) because our person measures were not well aligned with the item measures and the original calculation of PSR is affected by this misalignment.

The following iterations were performed: First, five items were rescored to scoring options [0 1 2 2 3] instead of [0 1 2 3 4] because of reversed thresholds. These five items were: Item 2 “*I avoid doing things that can expose me to the risk of hurting myself physically*”; item 3 “*I am perfectly aware of my physical limits*”; item 5 “*I trust that my body can learn new abilities*”; item 7 “*Doing vigorous physical activity can give me more strength and energy*”; and item 11 *“I do not feel comfortable pushing my body beyond its physical limits*”.

Next, item 11 (Fit Residual=6.99; *p*<0.0001), and item 2 (Fit Residual=6.33; *p*<0.0001) were deleted because of misfit. The misfit of item 11 is consistent with Menzel (2010) and Cossu *et al*. (2018)’s findings that this item was not associated with high loadings in their factor analysis. After deleting items 11 and 2, item 1 “*I do not feel ashamed of my body at all*” displayed misfit (Fit Residual=3.77, *p*=0.00016), but removing item 1 worsened the targeting (−1.89 ± 1.26) as well as the PSR value, which led us to the decision to keep this item in the scale. The resulting overall fit was χ^2^ (DF)=171.33 (80), p<0.0001. Table 2 shows the resulting item location in logits for each item after the above steps were completed.

**Table 2.**
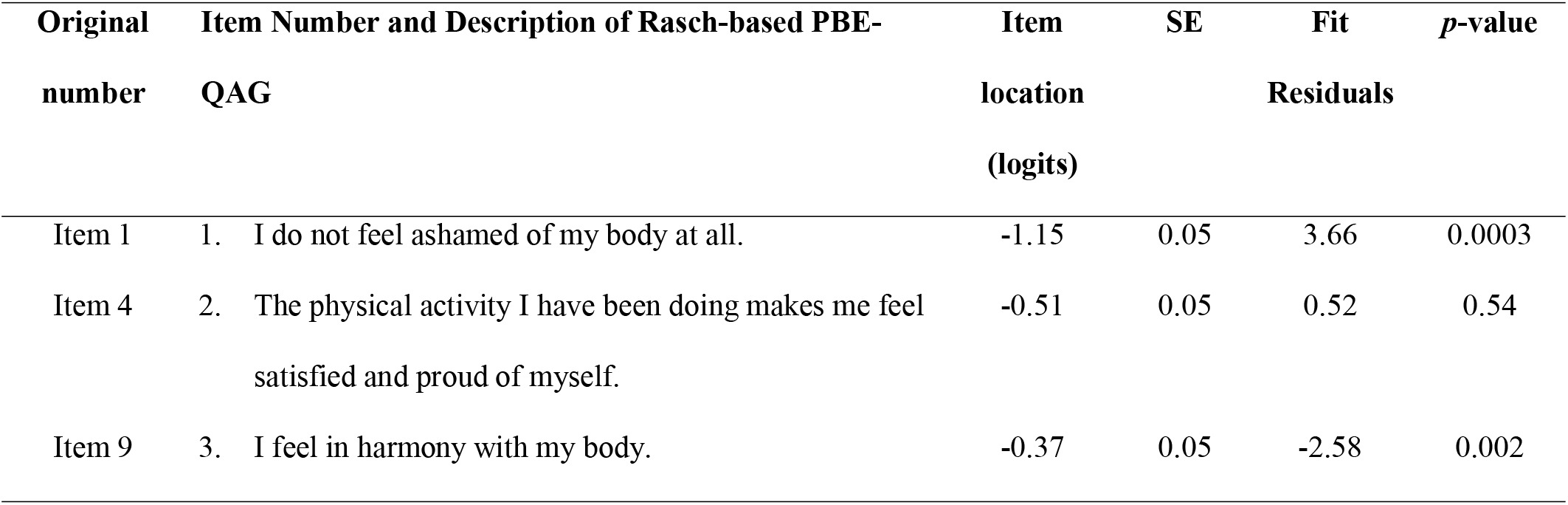
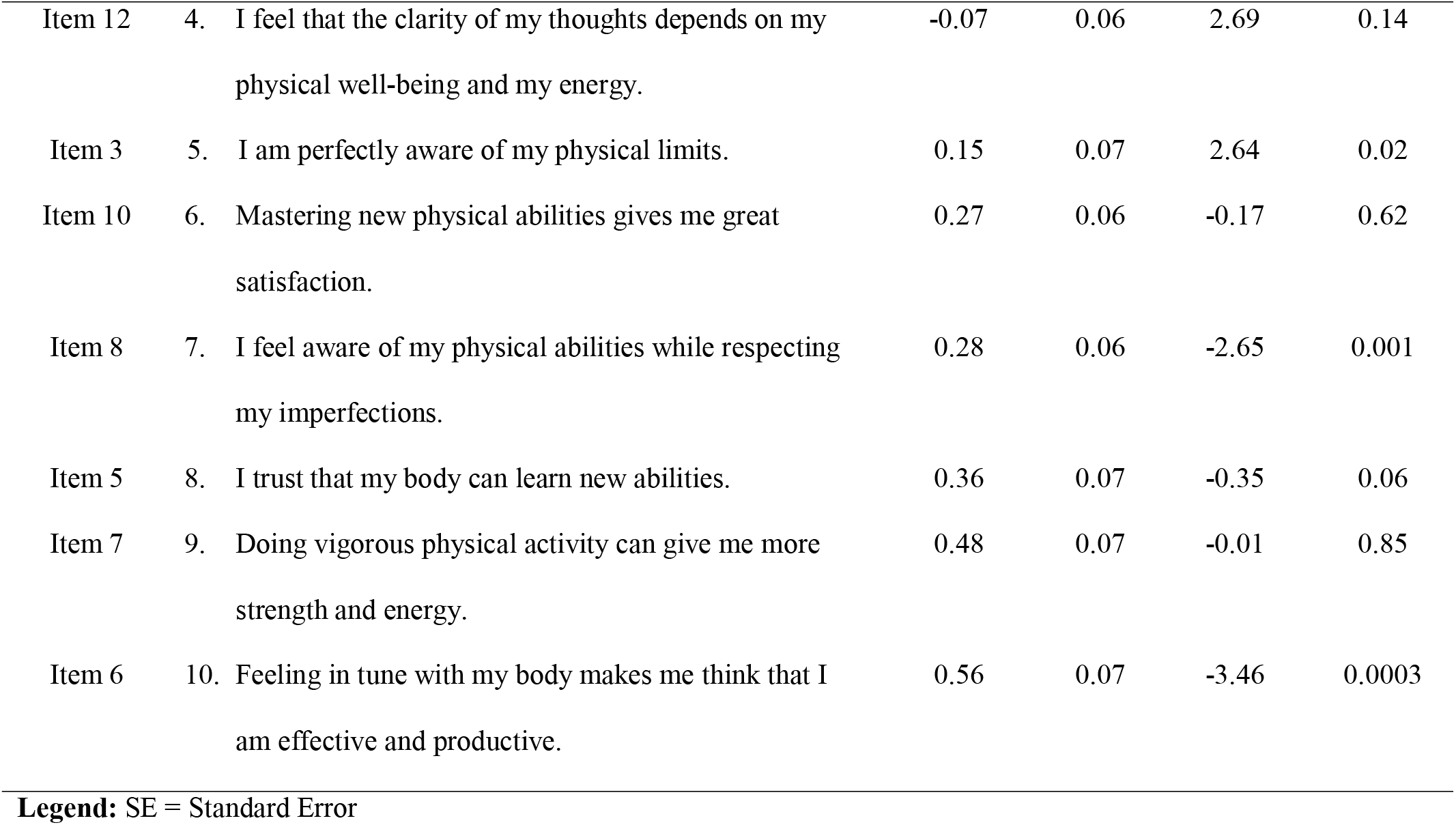
Item fit statistics of the Rasch-based PBE-QAG.

Table 3 shows the scoring categories of the Rasch-based BPE-QAG, taking into account the revisions proposed by the Rasch analysis.

**Table 3.**
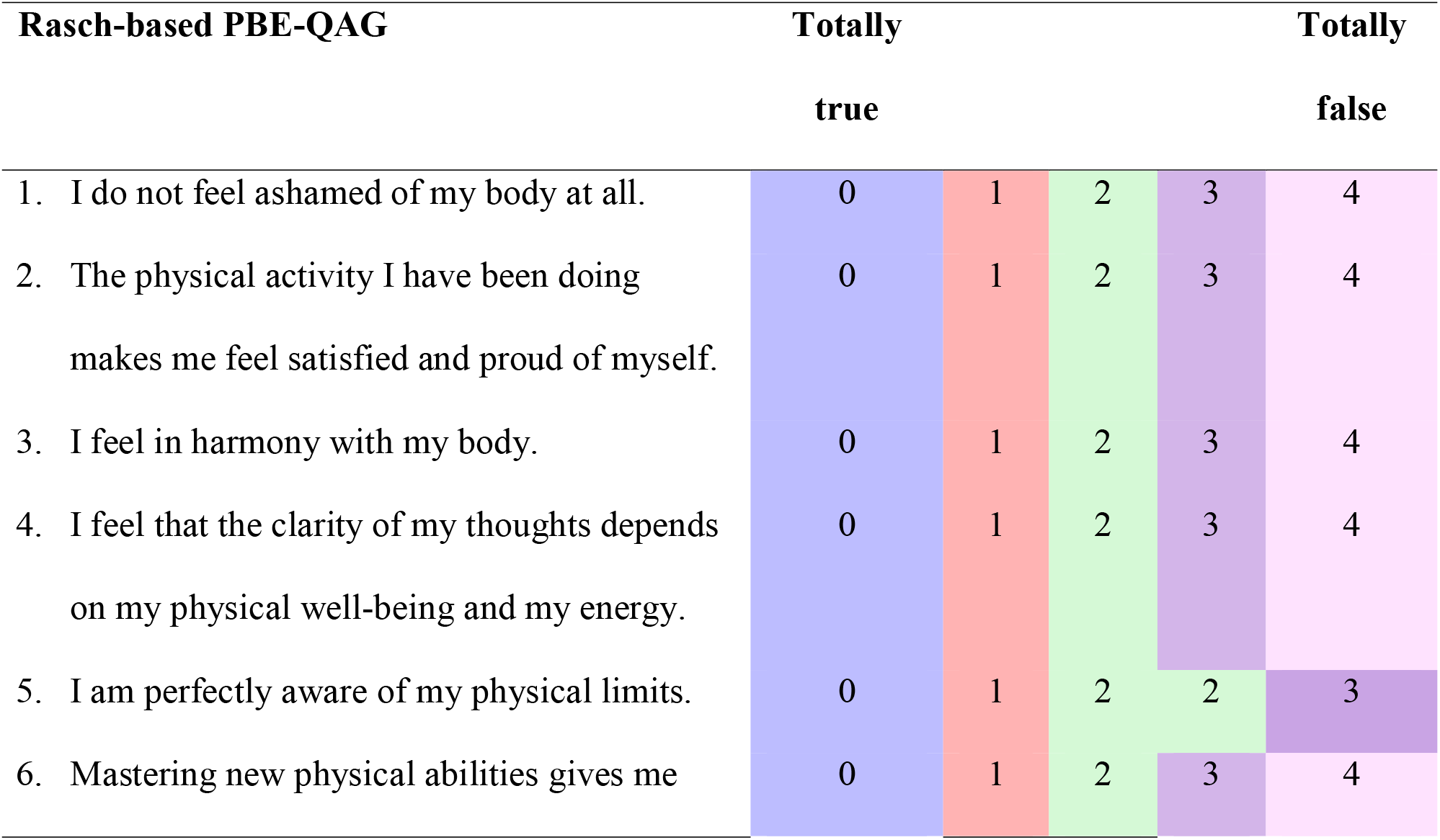
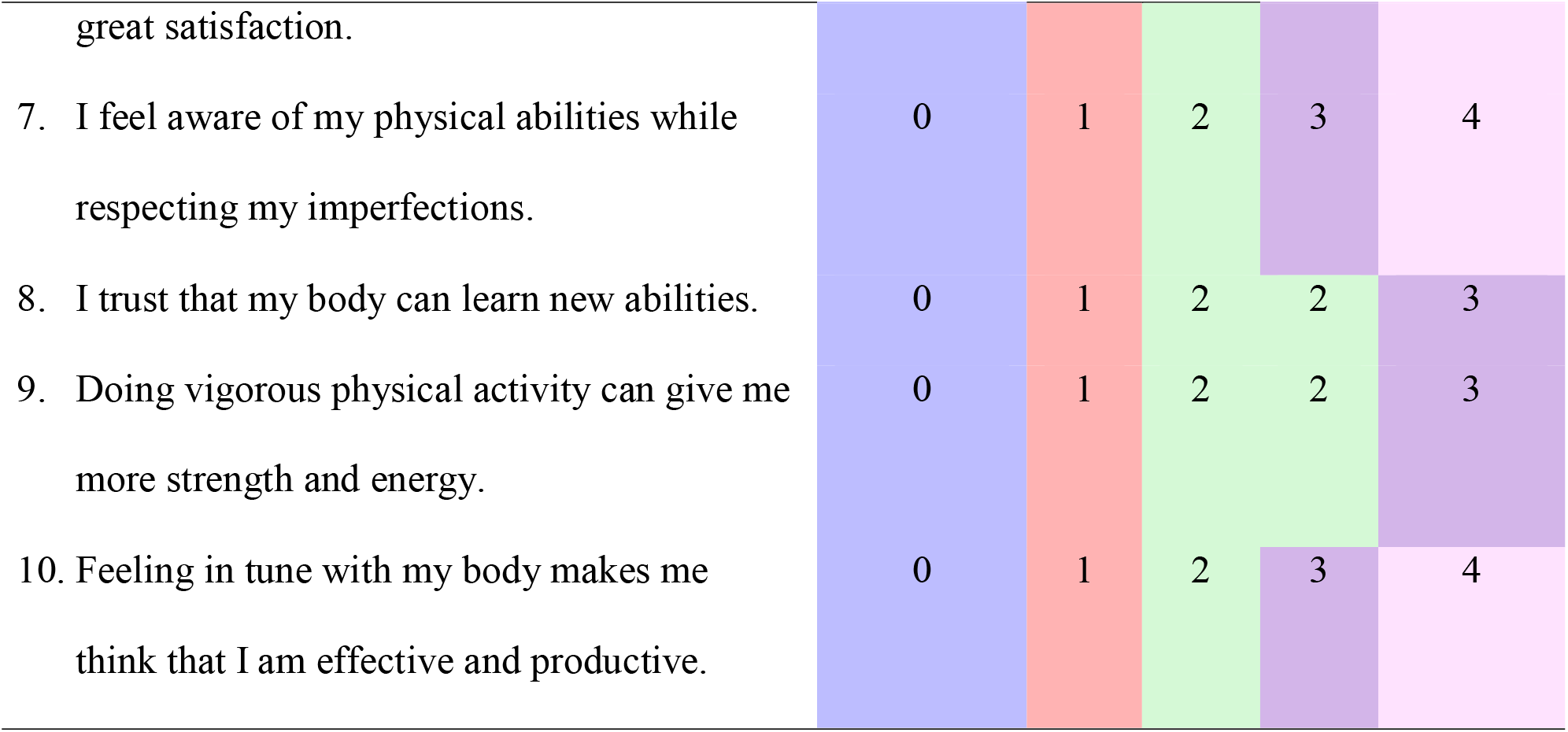
Revised Rasch-based BPE-QAG

Additionally, we found excellent person fit as only 1.23% (7 participants out of 566) had a misfit with a Fit Residual greater than 2.5. The item threshold map (Fig 1) displays the items hierarchy from the easiest item on the top to the hardest item on the bottom. There was minimal floor (5.72%) and no ceiling effect (0.00%). Reliability expressed through PSRw was high (0.96) with a mean standard error of 0.32 logits.

**Fig 1.**
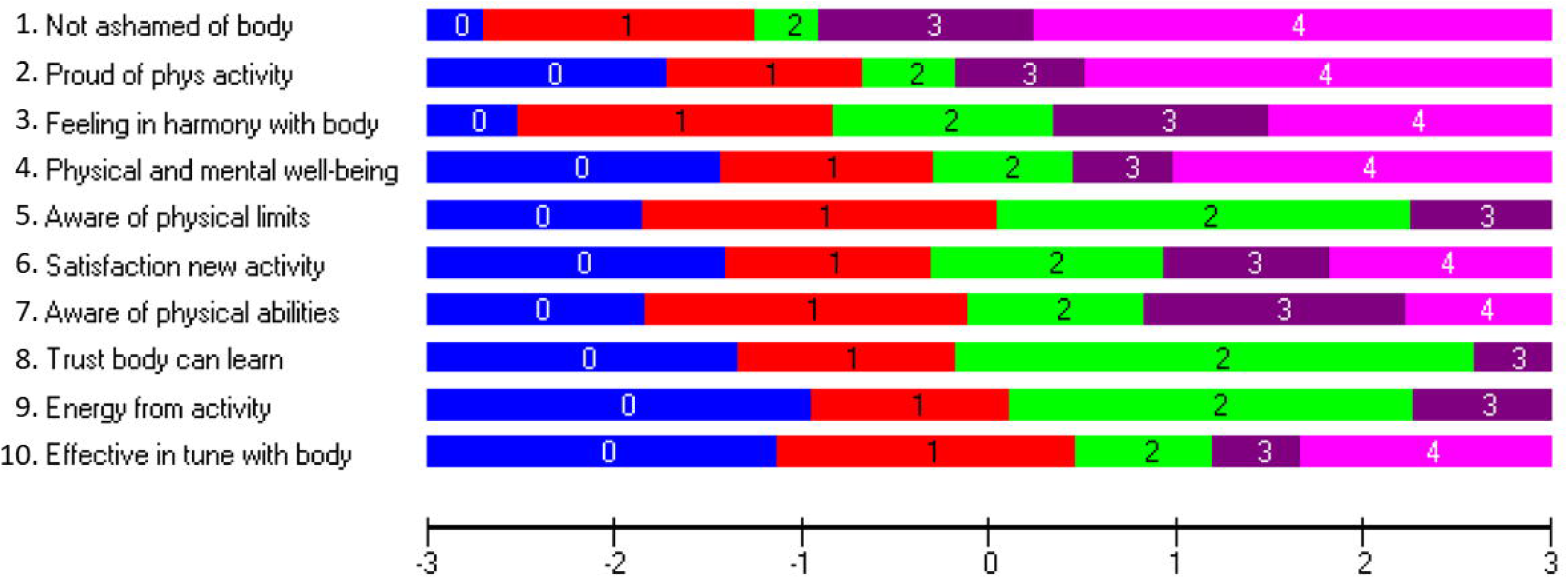
Item threshold map. The item threshold map shows the item hierarchy in terms of item difficulty, with the easiest item on top (original item number 1: *I do not feel ashamed of my body at all*) and the hardest item on the bottom (original item number 6: *Feeling in tune with my body makes me think that I am effective and productive*). The location of the thresholds between the scoring categories are represented on the logit scale at the bottom (the horizontal black line). This horizontal black line (interval logit scale) also represents the level of body awareness related to physical activity that participants have, with a higher score reflecting a lower body awareness level.

However, targeting of the person mean location relative to the item mean location was problematic: the person mean location was -1.71 ± 1.21 logits, as is demonstrated in Fig 2. Even though Cossu *et al*. (2018) designed the scale for older adults, our Rasch analysis showed that the average person location for adults who are 65 years old or older is -2.07±1.29, which demonstrates that the item difficulty of this scale is not well targeted for this age group. Additionally, the average person location for adults under 65 years of age is -1.59±1.15, which means that the targeting is off for this age group as well.

**Fig 2.**
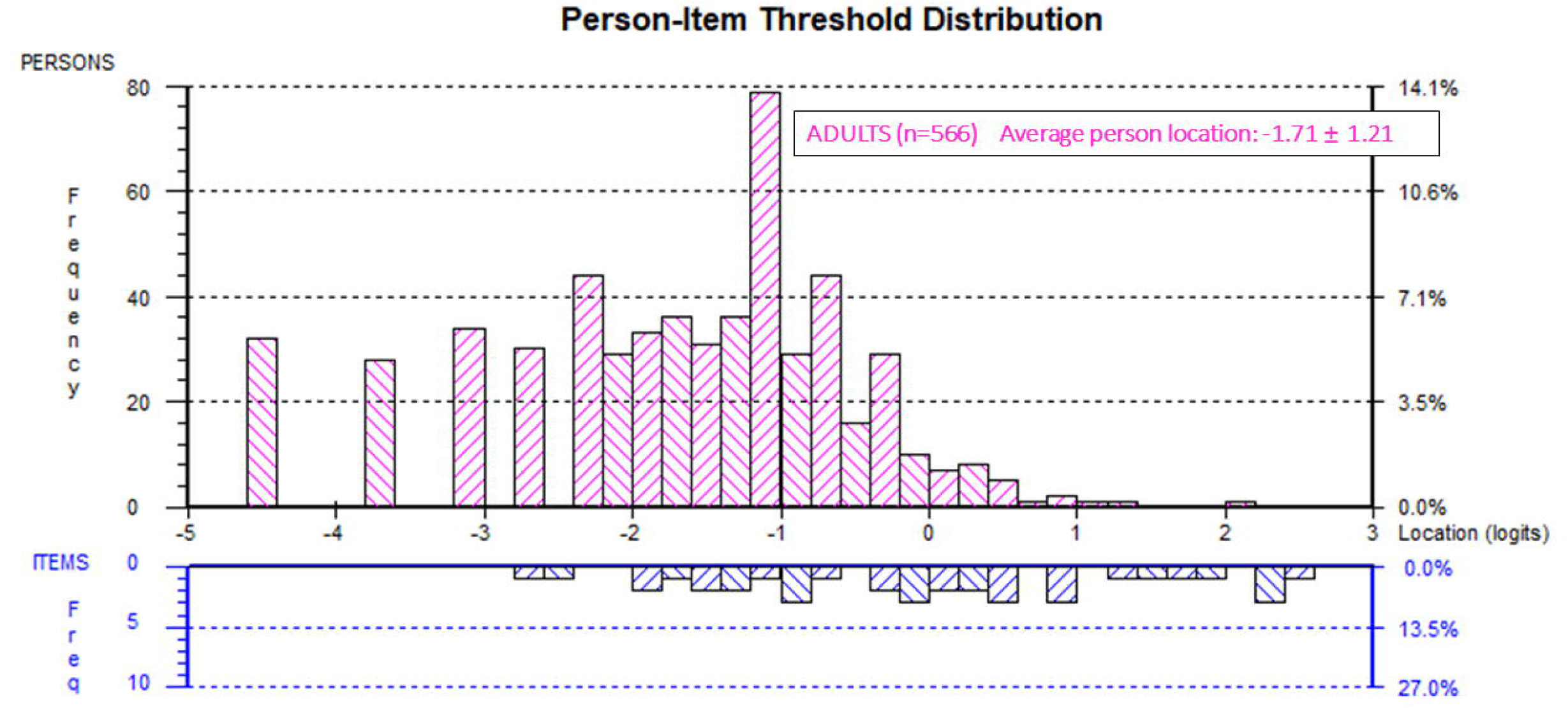
Person-Item threshold distribution. The pink histograms represent the frequency of participants at their body awareness ability level from a high level of body awareness (lowest logit value on the left side of the scale) to a low level of body awareness (highest logit value on the right side of the scale). The frequency of item thresholds is represented at the bottom of the figure (diagonal blue stripes histogram) on the same logit scale, organized from easiest items (left) to hardest items (right).

Furthermore, as can be seen in Table 4, the scoring categories 3 and 4 have the lowest response frequencies (i.e., less than 10 responses). This means that adults living in the community do not often report having low levels of body awareness related to physical activity.

**Table 4.** Frequency of categories

| Original<br>number | Item Number and Description of the Rasch-based PBE-QAG | 0<br>Totally<br>true | 1 | 2 | 3 | 4<br>Totally<br>false |
| --- | --- | --- | --- | --- | --- | --- |
| Item 1 | 1. I do not feel ashamed of my body at all | 90 | 172 | 133 | 105 | 34 |
| Item 4 | 2. The physical activity I have been doing makes<br>me feel satisfied and proud of myself | 199 | 171 | 98 | 49 | 17 |
| Item 9 | 3. I feel in harmony with my body | 140 | 212 | 133 | 42 | 7 |
| Item 12 | 4. I feel that the clarity of my thoughts depends<br>on my physical well-being and my energy | 240 | 184 | 81 | 24 | 5 |
| Item 3 | 5. I am perfectly aware of my physical limits | 207 | 246 | 77 | 4 |  |
| Item 10 | 6. Mastering new physical abilities gives me<br>great satisfaction | 250 | 181 | 83 | 17 | 3 |
| Item 8 | 7. I feel aware of my physical abilities while<br>respecting my imperfections | 215 | 214 | 83 | 19 | 3 |
| Item 5 | 8. I trust that my body can learn new abilities | 263 | 186 | 81 | 4 |  |
| Item 7 | 9. Doing vigorous physical activity can give me<br>more strength and energy | 308 | 165 | 57 | 4 |  |
| Item 6 | 10. Feeling in tune with my body makes me think<br>that I'm effective and productive | 297 | 180 | 45 | 9 | 3 |

There was no DIF greater than 0.50 logits for any of the variables. The PCAR analysis demonstrated an eigenvalue of the first contrast of 1.78 but the percent variance was 17.82%, which indicates there could be more than one dimension in this scale. Thus, there are subsets of items representing different aspects or dimensions of body awareness. The item subset with positive principal component loadings on the first contrast were items 5, 6, 7, and 10, likely reflecting a dimension of “awareness of physical skills”. The item subset with negative principal component loadings on the first contrast were items 1, 8 and 9, likely reflecting a dimension of “body acceptance”. The independent *t*-tests between the two subtests showed that only 3.94% of the independent *t*-tests were significant, thereby supporting the unidimensionality of the PBE-QAG.

There was no consequential LID as there were no standardized residual item correlations of at least 0.20 above the average residual item correlation (*r*=0.10).

Interestingly, we observed that 12 of the 26 adults with person measures above 0 logits had mental health conditions, such as anxiety, depression, or bipolar disorder. This led us to look at the person-item threshold distribution per group (Fig 3).

**Fig 3.**
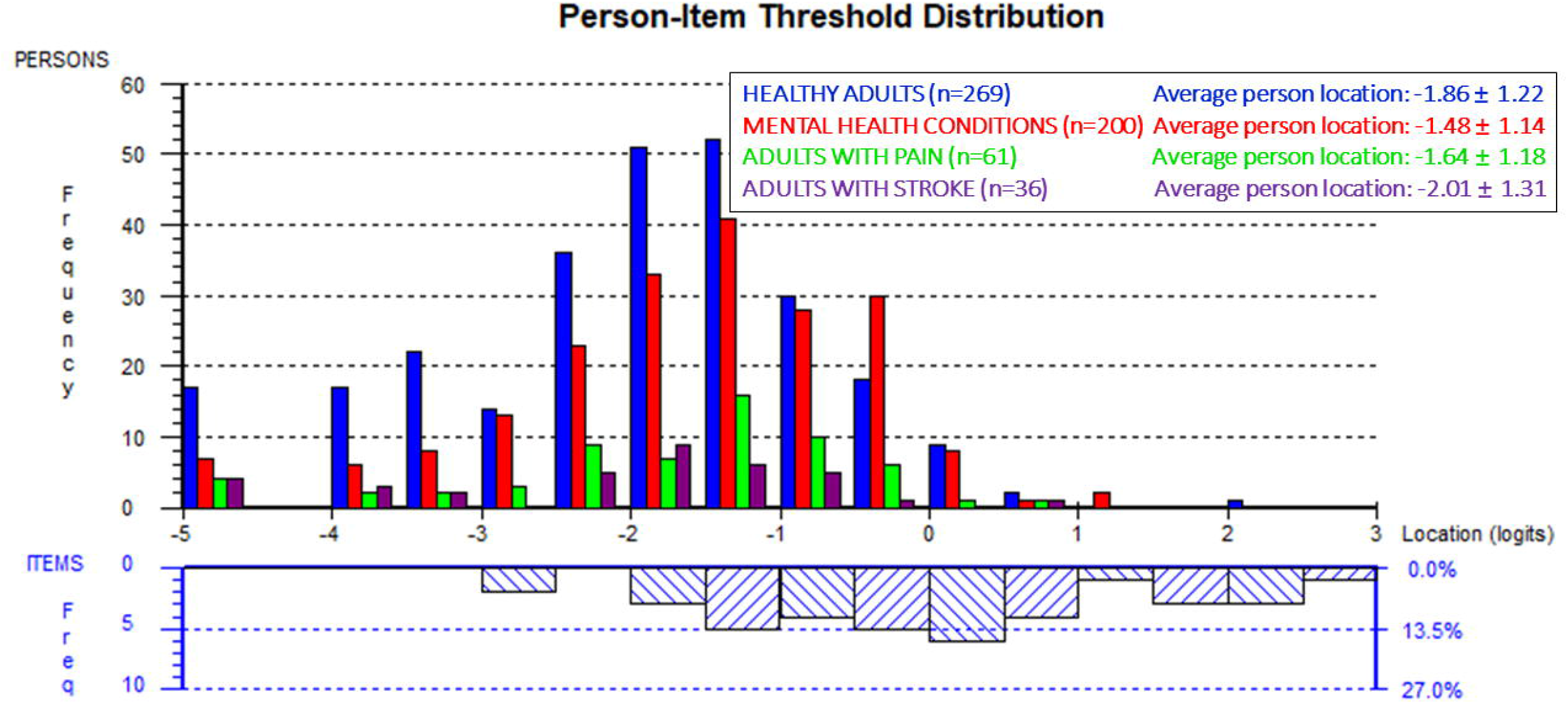
Person-Item threshold distribution per group. The pink histograms represent the frequency of the participants’ body awareness level (high body awareness level to low body awareness level, from left to right). The histograms at the top are organized per group, i.e., healthy adults (blue), adults with mental health conditions (red), adults with pain (green), and adults with stroke (purple). The frequency of item thresholds is represented at the bottom of the figure (diagonal blue stripes histogram) on the same logit scale, organized from easiest items (left) to hardest items (right).

Based on prior evidence of beneficial effects of body awareness training on mental health conditions,[43,44] we did a Kruskal-Wallis ANOVA test, to examine the impact on average person measures on four specific groups: healthy adults with and without currently performing body awareness training; and adults with mental health conditions with and without currently performing body awareness training. The result showed that adults with mental health conditions who are not doing any body awareness training have a significantly higher average person measure location (Median (IQR)=0.83(0.89) logits, *p*<0.0001)) compared to the other three groups (Fig 4).

**Fig 4.**
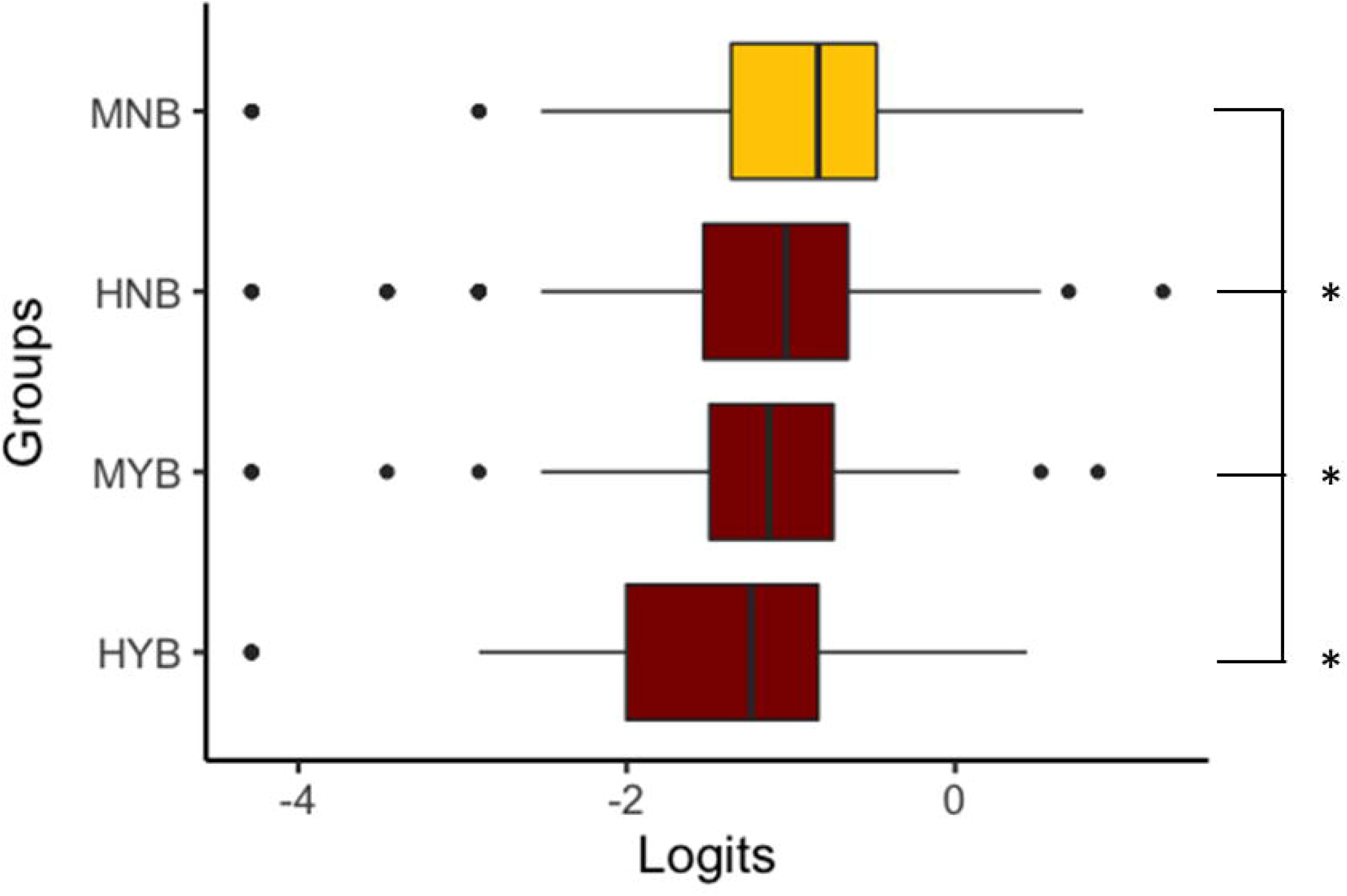
Kruskal-Wallis ANOVA for four groups divisions. HNB: healthy adults who are not doing any body awareness training. HYB: healthy adults who are doing body awareness training. MNB: adults with mental health conditions who are not doing any body awareness training. MYB: adults with mental health conditions who are doing body awareness training.

## Discussion

The present study demonstrates a Rasch-based validation of the PBE-QAG scale, which measures body awareness during physical activity. In relation to our aims, i.e., to evaluate the structural validity of the PBE-QAG with Rasch Measurement Theory in (i) healthy adults, to know which score range would be considered ‘normal’ body awareness related to physical activity; and in (ii) adults with known body awareness deficits, such as adults with mental health conditions, with pain, or with stroke, to verify whether this scale can accurately measure body awareness deficits in these populations, we present our two main findings below.

First, even though the Rasch validation reported a good item and person fit, the targeting is off for the adults recruited in our study. The average person location is -1.71±1.21, which means that participants in our study have a higher body awareness level than the PBE-QAG scale is designed to target. This means that adults living in the community would thus need a scale with more difficult items so that we can more accurately measure their level of body awareness related to physical activity. Therefore, the current form of the scale is not recommended for use in adults living in the community. Furthermore, the targeting of the scale would need to be solved before interpretations can be made about what a ‘normal scoring range’ of body awareness related to physical activity represents in healthy adults.

Because the targeting was off, we were also not able to clearly distinguish healthy adults from adults with pain or with stroke. Yet, we were able to demonstrate that adults with mental health conditions who do not perform body awareness training have a significantly higher average person measure location on the PBE-QAG than the adults who perform body awareness training or those who did not have mental health conditions. This leads us to presume that the current Rasch-based version of the scale might be more suited for adults with mental health conditions who are not currently training body awareness. Further validation of the scale will demonstrate if targeting can be achieved in this specific group. If so, this scale could then be used to evaluate changes in body awareness in adults with mental health conditions, after interventions that are known to improve body awareness, such as specific physical exercises or mind and body practices.

## Study limitations

Although we obtained a large sample size overall, our sample size of adults with stroke and adults with pain were smaller. Therefore, once items have been added to improve the targeting of the scale, a new Rasch validation in a larger sample of adults with stroke and adults with pain would be needed to ascertain the targeting and structural validity of the PBE-QAG in adults with stroke or pain.

## Conclusions

The Rasch-based PBE-QAG demonstrates good item and person fit, but the targeting is off. Adding items that target more complex aspects of body awareness related to physical activity might be needed before a new Rasch validation can be performed (i) to test whether the PBE-QAG is able to produce adequate targeting in people living in the community and (ii) to establish what a ‘normal range’ would represent in healthy adults. Based on our finding in our large sample size, we suggest that the current version of the Rasch-based PBE-QAG might be better suited for adults with mental health conditions who are not doing any body awareness training, but this needs to be validated with future studies.

## Data Availability

The raw data are deposited in the repository DRUM (University Digital Conservancy).

https://doi.org/10.13020/e6e1-as21

## Acknowledgments

The authors thank all participants as well as the volunteers who assisted the principal investigator (AVDW) at the Minnesota State Fair and at Highland Fest.

## Abbreviations

DIF: Differential Item Functioning
LID: Local Item Dependence
PBE-QAG: Physical Body Experiences Questionnaire Simplified for Active Aging
PCAR: Principal Components Analysis of Residuals
PSR: Person Separation Reliability
RUMM2030: Rasch Unidimensional Measurement Model 2030 software

## Supporting information

**S1 Table. Iterations analysis table of the PBE-QAG**. DF=Degrees of Freedom; PCAR=Principal Components Analysis of Residuals; PSR=Person Separation Reliability; PSRw=Wright corrected PSR; RMSE=Root Mean Square Error (reflected as Mean Error Variance in RUMM2030)

