## Supplementary material for "Physical body experiences questionnaire simplified for active aging (PBE-QAG): Validation with Rasch measurement theory": S1 Table

| **Analysis** | **Items** | **Rating scale categories** | **Person mean (SD) logits** | **Mean error variance** | **Floor effect**  **n (%)** | **Ceiling effect**  **n (%)** | **Overall Chi-square (DF)**  **p-value** | **PSR**  **(PSRw)** | **Items with disordered thresholds**  **(n)** | **Misfitting items**  **(n)** | **PCAR**  **Eigenvalue**  **1^st^ contrast**  **(%)** | **Misfitting persons**  **n (%)** |
| --- | --- | --- | --- | --- | --- | --- | --- | --- | --- | --- | --- | --- |
| All items  (n=566) | 12 | 60 | -1.19  (0.87) | 0.17 | 13  (2.30%) | 0  (0%) | 569.54  (96)  *p*<0.0001 | 0.74  (0.97) | 5 | 2  (items 2, 11) | 2.43 (20.27%) | 9 (1.59%) |
| 5 items rescored to 0 1 2 2 3  (n=566) | 12 | 55 | -1.30 (0.94) | 0.19 | 13  (2.30%) | 0  (0%) | 416.11  (96)  *p*<0.0001 | 0.76  (0.97) | 0 | 2  (items 2, 11) | 2.17  (18.07%) | 9 (1.59%) |
| Item 11 deleted (n=566) | 11 | 51 | -1.48  (1.05) | 0.24 | 18 (3.18%) | 0  (0%) | 361.35 (88)  *p*<0.0001 | 0.77 (0.97) | 0 | 1  (item 2) | 1.79  (16.28%) | 8 (1.41%) |
| Item 2 deleted (n=566) | 10 | 47 | -1.71 (1.21) | 0.32 | 32 (5.65%) | 0  (0%) | 171.33  (80)  *p*<0.0001 | 0.77 (0.96) | 0 | 1  (item 1) | 1.78 (17.82%) | 8 (1.41%) |
